# Change in Effectiveness of Sotrovimab for Preventing Hospitalization and Mortality in COVID-19 Outpatients During the Omicron Phase

**DOI:** 10.1101/2022.06.17.22276575

**Authors:** Neil R. Aggarwal, Laurel E. Beaty, Tellen D. Bennett, Nichole E. Carlson, Adit A. Ginde

**Affiliations:** Department of Medicine, University of Colorado School of Medicine, Aurora, 80045, USA; Department of Biostatistics and Informatics, Colorado School of Public Health, Aurora, 80045, USA; Section of Informatics and Data Science, Department of Pediatrics, University of Colorado School of Medicine, Aurora, 80045, USA; Department of Emergency Medicine, University of Colorado School of Medicine, Aurora, 80045, USA; Colorado Clinical and Translational Sciences Institute, University of Colorado Anschutz Medical Campus, Aurora, 80045, USA

**Keywords:** real-world evidence, COVID-19, sotrovimab, outpatients, mortality

## Abstract

**Background:** Sotrovimab, a neutralizing monoclonal antibody (mAb) treatment authorized for early symptomatic COVID-19 patients, was effective in preventing the progression of severe disease and mortality following SARS-CoV-2 Delta variant infection. It is not known whether sotrovimab is similarly effective for SARS-CoV-2 Omicron variant infection.

**Methods:** Observational cohort study of non-hospitalized adult patients with SARS-CoV-2 infection from December 26, 2021 to March 10, 2022 (>96% Omicron BA.1 variant), using electronic health records from a statewide health system linked to state-level vaccine and mortality data. We used propensity matching to select up to 3 patients not receiving mAbs or other authorized antivirals for each patient who received outpatient sotrovimab treatment. The primary outcome was 28-day hospitalization; secondary outcomes included mortality. To evaluate change in sotrovimab effectiveness during the Omicron phase, we propensity matched sotrovimab-treated patients from Omicron to Delta (October 1-December 11, 2021) phases to each other and then to untreated controls with a treatment-variant interaction added to the logistic regression model.

**Results:** Of 30,247 patients with SARS-CoV-2 infection, we matched 1,542 receiving sotrovimab to 3,663 not receiving treatment. Compared to untreated patients, sotrovimab treatment was not associated with reduced odds of all-cause 28-day hospitalization (raw rate 2.5% versus 3.2%; adjusted OR 0.82, 95% CI 0.55, 1.19) or mortality (raw rate 0.1% versus 0.2%; adjusted OR 0.62, 95% CI 0.07, 2.78). In the combined analysis across Omicron and Delta phases, the observed treatment OR was higher during Omicron than during Delta (OR 0.85 vs. 0.39, respectively; interaction p=0.053)

**Conclusion:** Real-world evidence demonstrated sotrovimab was not associated with reduced hospitalization and all-cause 28-day mortality among COVID-19 outpatients during the Omicron BA.1 phase and attenuated compared to the Delta phase

**Summary:** Real-world evidence demonstrates that the neutralizing monoclonal antibody sotrovimab was not associated with lower 28-day hospitalization and mortality rates when administered to high-risk outpatients recently infected with SARS-CoV-2 during the Omicron variant phase, compared to a propensity-matched cohort of untreated outpatients.

## BACKGROUND

With fluctuating rates of transmission of severe acute respiratory syndrome coronavirus-2 (SARS-CoV-2), readily available neutralizing monoclonal antibody (mAb) products such as sotrovimab for outpatients who have recently tested positive for SARS-CoV-2 has been a critical, evidence-based treatment strategy to mitigate the impact of COVID-19 surges on the health care system and improve COVID-19 outcomes among high-risk individuals.[1-7] Several mAb products received emergency use authorization (EUA) from the US Food and Drug Administration[8] based on Phase II/III randomized clinical trials conducted earlier in the pandemic that demonstrated efficacy towards reduced hospitalization and disease severity among high-risk outpatients,[9-11] but little randomized data is available to inform mAb efficacy against new variants including Omicron lineages that have rapidly evolved.[12] As such, analysis of more contemporaneous real-world data sufficiently robust to evaluate important clinical differences is critical to evaluate treatment effectiveness and inform policy and practice decisions, as we and others have successfully done.[3-7]

We previously used a real-world data platform to report on sotrovimab effectiveness during the Delta variant pandemic phase,[2] adding to the evidence generated from the COMET-ICE trial that found a significant reduction in risk of a composite endpoint of all-cause hospitalization or death following sotrovimab treatment [10] and led to emergency use authorization (EUA) for sotrovimab (May 26, 2021). Subsequently, when EUAs for other authorized mAbs were revoked in January 2022, sotrovimab was the only available mAb for outpatient treatment during early Omicron. However, a marked reduction in sotrovimab *in vitro* neutralization against Omicron BA.2 and its sublineages [13, 14] led to the sotrovimab EUA being fully revoked when Omicron BA.2 sub-variant prevalence was estimated to be greater than 50% in all HHS U.S. regions (April 5, 2022). In support of this determination, a report of real-world clinical data suggested sotrovimab ineffectiveness in reducing rates of COVID-19 progression during BA.2 Omicron subvariant dominant phase in Qatar.[15] However, a recent report in immunocompromised solid organ transplant recipients suggested a benefit of sotrovimab in reducing severity of illness following early administration after SARS-CoV-2 infection during the Omicron BA.1 phase,[16] yet sotrovimab evaluation of effectiveness against Omicron BA.1 or BA.1.1 in a broader population of high-risk outpatients is lacking.

To provide additional data on sotrovimab effectiveness against Omicron SARS-CoV-2, including immunocompromised patients and other higher-risk subgroups, we used our real-world data platform to assess the impact of sotrovimab treatment on hospitalization and mortality among outpatients with early symptomatic COVID-19 infections during a SARS-CoV-2 Omicron BA.1 predominant phase in Colorado (December 26, 2021to March 10, 2022).[17-20]

## METHODS

### Study Oversight and Data Sources

We conducted a propensity-matched observational cohort study, as part of a statewide implementation/effectiveness pragmatic trial, in a collaboration between University of Colorado researchers, University of Colorado Health (UCHealth) system leaders, and the Colorado Department of Public Health and Environment (CDPHE). The study was approved by the Colorado Multiple Institutional Review Board with a waiver of informed consent. We obtained data from the electronic health record (EHR; Epic, Verona, WI) of UCHealth, the largest health system in Colorado with 13 hospitals around the state and 141,000 annual hospital admissions, using Health Data Compass, an enterprise-wide data warehouse. EHR data were merged with statewide data on vaccination status from the Colorado Comprehensive Immunization Information System and mortality from Colorado Vital Records.

### Patient Population

Our primary cohort was patients who were diagnosed with SARS-CoV-2 infection between December 26, 2021 and March 10, 2022. Based on Colorado state-wide data [17] SARS-CoV-2 infections due to the Omicron variant made up at least 96% of overall cases by 12/26/2021. As an update on previously published work,[2] a second cohort was selected between October 1, 2021 to December 11, 2021 when the Delta variant made up at least 99% of overall cases to be able to investigate potential changes in effectiveness between the Delta and Omicron phases. All patients had least 28 days of follow-up. Patients were identified by either a positive SARS-CoV-2 test (by polymerase chain reaction or antigen) or by the date of mAb administration if the date of the SARS-CoV-2 positive test was missing.

For our primary cohort, we excluded patients who received a medication order for any anti-viral treatment except sotrovimab within 10 days of the positive SARS-CoV-2 test (**Appendix Figure 1**); thereby we included only patients who were untreated (N = 31,187) or who were treated with sotrovimab (N = 1,683). We excluded patients who were missing both a positive SARS-CoV-2 test date and a mAb administration date (N = 605), those who were already in the hospital or who were hospitalized on the same day as the positive test (N = 2,009), and if more than 10 days had elapsed between the SARS-CoV-2 test and mAb administration (N = 9). We did not exclude patients based on EUA eligibility due to the lack of consistently available comprehensive EHR data for all patients. After exclusions, the cohort included 28,584 untreated patients and 1,663 sotrovimab-treated patients. We applied the same exclusions to the Delta cohort, resulting in 8,901 untreated patients and 556 sotrovimab-treated patients (**Appendix Figure 2**).

### Outcomes

The primary outcome was all-cause hospitalization within 28-days of the SARS-CoV-2 positive test. Secondary outcomes included all-cause 28-day mortality and 28-day Emergency Department (ED) visit were also computed. For both hospitalization and ED visit, we used the index visit. For patients that were hospitalized, we evaluated disease severity based on the maximum level of respiratory support required, intensive care unit (ICU) admission rate, hospital and ICU length of stay (LOS) in survivors, and in-hospital mortality.

### Variable Definitions

We used EHR data to identify all outcomes of interest. A hospitalization was defined as any inpatient or observation encounter and ED visits were defined as any visit to the ED, with or without an associated hospitalization. We estimated disease severity on an ordinal scale with the maximum level of respiratory support used at an encounter level with the following possible types (in ascending order): no supplemental oxygen, standard (nasal cannula/face mask) oxygen, high-flow nasal cannula (HFNC) or non-invasive ventilation (NIV), and invasive mechanical ventilation (IMV). In-hospital mortality was the highest level of disease severity. Due to small sample sizes, we also categorized disease severity as HFNC/NIV/IMV/Death vs. standard oxygen/no supplemental oxygen.

We determined presence and status of comorbid conditions based on previously described methods using the Charlson and Elixhauser Comorbidity Indices.[2, 20] Immunocompromised status was categorized as not immunocompromised, mild immunocompromised, and moderately/severe immunocompromised based on pre-hospitalization use of immunosuppressive medications and immunosuppressive conditions (**Appendix Table 1**). We calculated the total number of comorbidities as the sum of the presence of diabetes mellitus, cardiovascular disease, pulmonary disease, renal disease, hypertension, and liver disease and classified as none, one, or two or more; we analyzed immunocompromised status and obesity as separate variables. We categorized vaccination status as the total number of vaccinations (0, 1, 2, or ≥ 3) administered prior to the date of the SARS-CoV-2 positive test.

Other variables of interest include treatment status, categorical age in years, sex, race/ethnicity, insurance status, obesity status, immunocompromised status, number of comorbidities, vaccination status, and cohort week (**Table 1, 2**). In the statistical models, Medicare and private/commercial were collapsed into one category due to collinearity of Medicare with age.

**Table 1.** Baseline Characteristics by Mab Treatment Status for Primary Matched Cohort.

|  | Untreated N=3663) | Sotrovimab Treated (N=1542) |
| --- | --- | --- |
| **Age Group \*** |  |  |
| 18-44 years | 997 (27.2%) | 387 (25.1%) |
| 45-64 years | 1219 (33.3%) | 492 (31.9%) |
| ≥65 years | 1447 (39.5%) | 663 (43.0%) |
| **Female Gender \*** | 2231 (60.9%) | 916 (59.4%) |
| **Race/Ethnicity \*** |  |  |
| Non-Hispanic White | 2974 (81.2%) | 1256 (81.5%) |
| Hispanic | 391 (10.7%) | 160 (10.4%) |
| Non-Hispanic Black | 95 (2.6%) | 41 (2.7%) |
| Other | 203 (5.5%) | 85 (5.5%) |
| **Insurance Status \*** |  |  |
| Private/Commercial | 1878 (51.3%) | 759 (49.2%) |
| Medicare | 1581 (43.2%) | 699 (45.3%) |
| Medicaid | 110 (3.0%) | 48 (3.1%) |
| None/Uninsured | 3 (0.1%) | 2 (0.1%) |
| Other/Unknown | 91 (2.5%) | 34 (2.2%) |
| **Immunocompromised Status \*** |  |  |
| None | 2148 (58.6%) | 902 (58.5%) |
| Mild | 679 (18.5%) | 275 (17.8%) |
| Moderate/Severe | 836 (22.8%) | 365 (23.7%) |
| **Obesity\*** | 1119 (30.5%) | 483 (31.3%) |
| **Number of Other Comorbid Conditions \*** |  |  |
| None | 998 (27.0%) | 411 (26.7%) |
| One | 914 (25.0%) | 393 (25.5%) |
| Two or more | 1761 (48.1%) | 738 (47.9%) |
| **Diabetes Mellitus** | 716 (19.5%) | 371 (24.1%) |
| **Cardiovascular Disease** | 1116 (30.5%) | 446 (28.9%) |
| **Pulmonary Disease** | 1295 (35.4%) | 540 (35.0%) |
| **Renal Disease** | 446 (12.2%) | 268 (17.4%) |
| **Hypertension** | 1812 (49.5%) | 765 (49.6%) |
| **Liver Disease** | 488 (13.3%) | 235 (15.2%) |
| **Number of vaccinations prior to SARS-CoV-2+ \*** |  |  |
| 0 | 898 (24.5%) | 335 (21.7%) |
| 1 | 182 (5.0%) | 68 (4.4%) |
| 2 | 726 (19.8%) | 284 (18.4%) |
| 3+ | 1857 (50.7%) | 855 (55.4%) |
| **Days to mAb Admin: mean (SD)** | NA | 3.004 (1.967) |
| **Time (weeks)\*** |  |  |
| 12/26 - 1/1 | 704 (19.2%) | 258 (16.7%) |
| 1/2 - 1/8 | 353 (9.6%) | 106 (6.9%) |
| 1/9 - 1/15 | 382 (10.4%) | 127 (8.2%) |
| 1/16 - 1/22 | 500 (13.7%) | 170 (11.0%) |
| 1/23 - 1/29 | 393 (10.7%) | 128 (8.3%) |
| 1/30 - 2/5 | 442 (12.1%) | 188 (12.2%) |
| 2/6 - 2/12 | 291 (7.9%) | 167 (10.8%) |
| 2/13 - 2/19 | 211 (5.8%) | 112 (7.3%) |
| 2/20 - 2/26 | 181 (4.9%) | 107 (6.9%) |
| 2/27 - 3/5 | 116 (3.2%) | 102 (6.6%) |
| 3/6 - 3/10 | 90 (2.5%) | 77 (5.0%) |

<sup>a</sup> Variables used in the propensity matching. Abbreviations: mAb, monoclonal antibody.

**Table 2.** Primary and Secondary Outcomes by Monoclonal Antibody Treatment Status for Primary Cohort.

| **Outcome** | **Sotrovimab-Treated** | **Untreated** | **Adjusted OR** | **95% CI** |
| --- | --- | --- | --- | --- |
| **Overall Sample Size** | **N=1542** | **N=3663** |  |  |
| All-Cause 28-day Hospitalization<br>(*primary outcome*) | 39 (2.5%) | 116 (3.2%) | 0.82 | (0.55, 1.19) |
| All-Cause 28-day Mortality | 1 (0.1%) | 7 (0.2%) | 0.62 | (0.07, 2.78) |
| Any ED visit to Day 28 | 93 (6.0%) | 224 (6.1%) | 1.03 | (0.79, 1.32) |
| **Hospitalized Sample Size** | **N=39** | **N=116** |  |  |
| Hospital LOS days, mean (SD) | 5.2 (6.8) | 7.3 (7.7) | -- | -- |
| HFNC/NIV, IMV or Death | 6 (15.4%) | 33 (28.4%) | -- | -- |
| ICU Admission | 4 (10.3%) | 20 (17.2%) | -- | -- |
| ICU LOS days, mean (SD) | 1.5 (0.58) | 8.1 (14.04) | -- | -- |

All regression models adjusted for age, sex, race/ethnicity, obesity, immunocompromised status, number of comorbidities, insurance status, and vaccination status. Abbreviations: mAb, monoclonal antibody; OR, odds ratio; CI, confidence interval; ICU, intensive care unit; IMV, invasive mechanical ventilation; LOS, length of stay; SD, standard deviation; HFNC, high flow nasal cannula; NIV, non-invasive ventilation

### Statistical Analysis

#### Omicron only analysis

We used nearest neighbor propensity matching with logistic regression to match patients with treatment status as the outcome. The propensity model included age, sex, race/ethnicity, insurance status, obesity status, immunocompromised status, number of other comorbid conditions, number of vaccinations, and week in the study (categorical). We removed 67 sotrovimab treated patients who had missing covariate data and lost an additional 54 in the matching process. We assessed the achieved balance using a threshold of <0.1 for the standardized mean differences (SMD) and achieved a ratio of 2.38:1 (3,663:1,542) untreated to treated patients. As previously,[2] patients who were missing a SARS-CoV-2 positive test date (56.9%) had their test date randomly imputed test based on the distribution of observed time to mAb treatment.

We used Firth’s logistic regression to assess associations between binary outcomes (28-day hospitalization, 28-day mortality, and 28-day ED visits) and treatment. Firth’s logistic regression (R package logistf V 1.24) addresses the issues with low event rates and complete separation.[21] Each multivariable model included all variables of interest as outlined in the previous section. We included cohort week as a continuous, linear term in all adjusted models and constructed cumulative incidence curves to visually assess the trend across time from SARS-CoV-2 positive date to 28-day hospitalization by treatment status. We also analyzed in-hospital secondary outcomes related to severity of respiratory disease in a descriptive manner.

We focused on five subgroup analyses of clinical interest: age (<65 years vs. ≥65 years), immunocompromised status (both binary and tri-level), number of comorbid conditions (≥2 vs. <2), number of vaccinations (≥3 vs. <3), and time of study in the Omicron phase (Early vs. Late). The treatment effect for each subgroup was estimated using interaction models. Each model was adjusted for all variables included in the primary model.

Two sensitivity analyses were performed. We repeated the primary analysis including only patients that we were able to verify their EUA eligibility based on available EHR data. We also repeated the primary analysis using a different SARS-CoV-2 positive test date imputation method. The second method imputed a 10-day difference from the observed mAb administration date (the maximum difference allowed by the EUA).

#### Omicron and Delta analysis

To compare the effect of sotrovimab treatment during Omicron- or Delta-predominant COVID-19 phases, we developed a second propensity matched analysis cohort. First, to address imbalances in treatment cohorts due to supply, sotrovimab-treated patients during the Omicron-predominant phase were nearest-neighbor propensity matched to sotrovimab treated patients in Delta-predominant phase based on a logistic regression with variant as the outcome. Matching variables included age, sex, race/ethnicity, obesity, immunocompromised status, number of comorbid conditions, number of vaccinations, and insurance status. Then we propensity matched the above matched sotrovimab-treated patient to untreated patients stratified by variant using nearest-neighbor matching based on a logistic regression with treatment status as the outcome and the same covariates previously described.

We fit Firth’s logistic regression models with all-cause 28-day hospitalization as the outcome: 1) the model was stratified by variant and included all variables of interest from the primary analysis and 2) an analysis with both cohorts combined with a treatment-variant interaction added to the logistic regression model along with the adjustment variables. The second model allows us to formally test if the effect of treatment differed statistically between the Delta and Omicron cohorts.

All statistical analyses were performed using R Statistical Software (version 3.6.0; R Foundation for Statistical Computing).[22]

## RESULTS

### Characteristics of sotrovimab-treated and untreated cohorts in the primary cohort

Of 30,247 patients with SARS-CoV-2 infection in the full primary cohort, 1,663 subjects received mAbs and 28,584 patients did not (**Appendix Table 2**). In the full primary cohort, the sotrovimab-treated group generally reflects EUA criteria for use of mAbs. Those treated were older (44.3% were age ≥65 years vs. 11.4% in untreated group), more likely to be obese (30.5% vs. 16.5%), be immunocompromised at any severity level (40.5% vs. 12.9%) or have one or more comorbid conditions (71.9% vs. 37.6%). Particularly in early Omicron cohort weeks, the rate of sotrovimab treatment was lower as compared to the untreated group, due to a surge in cases relative to available treatment. Propensity matching eliminated clinically meaningful differences in matching variables between groups, resulting in 1,542 sotrovimab-treated patients propensity matched to 3,663 untreated patients (**Appendix Figure 1**).

The characteristics of sotrovimab-treated and untreated patients in the matched primary cohort are presented in **Table 1**. Overall, the age distribution was similar with approximately 40% aged ≥65 years, 60% female, 80% Non-Hispanic white, and 50% with private/commercial insurance. Hypertension (49%) and pulmonary disease (35%) were the most common comorbid conditions. Notably, 50.7% vs. 55.4% of untreated and sotrovimab-treated patients had received three or more vaccine doses at the time of infection, and 24.5% vs. 21.7% had not received any vaccine doses, respectively. The mean time from positive SARS-CoV-2 test to administration of sotrovimab treatment was 3.0 days (SD 1.8) in those with a positive test date in the EHR.

### Hospitalization and Mortality

Sotrovimab treatment was not associated with a lower rate of 28-day hospitalization compared to matched untreated controls ((39 [2.5%] vs. 116 [3.2%]), adjusted OR (aOR) = 0.82 (95% CI 0.55-1.19; p = 0.29)) (**Table 2, Figure 1**). Covariates that were associated with increased odds of 28-day hospitalization included age ≥ 65 (p = 0.04), obesity (p = 0.02), moderate/severe immunocompromised status (p < 0.001), and two or more other comorbid conditions (p < 0.001) (**Appendix Table 3, Supplement**). Having received two (p = 0.03) or ≥ three (p < 0.001) vaccine doses were both associated with reduced hospitalization in comparison to having zero vaccine doses.

**Figure 1.**
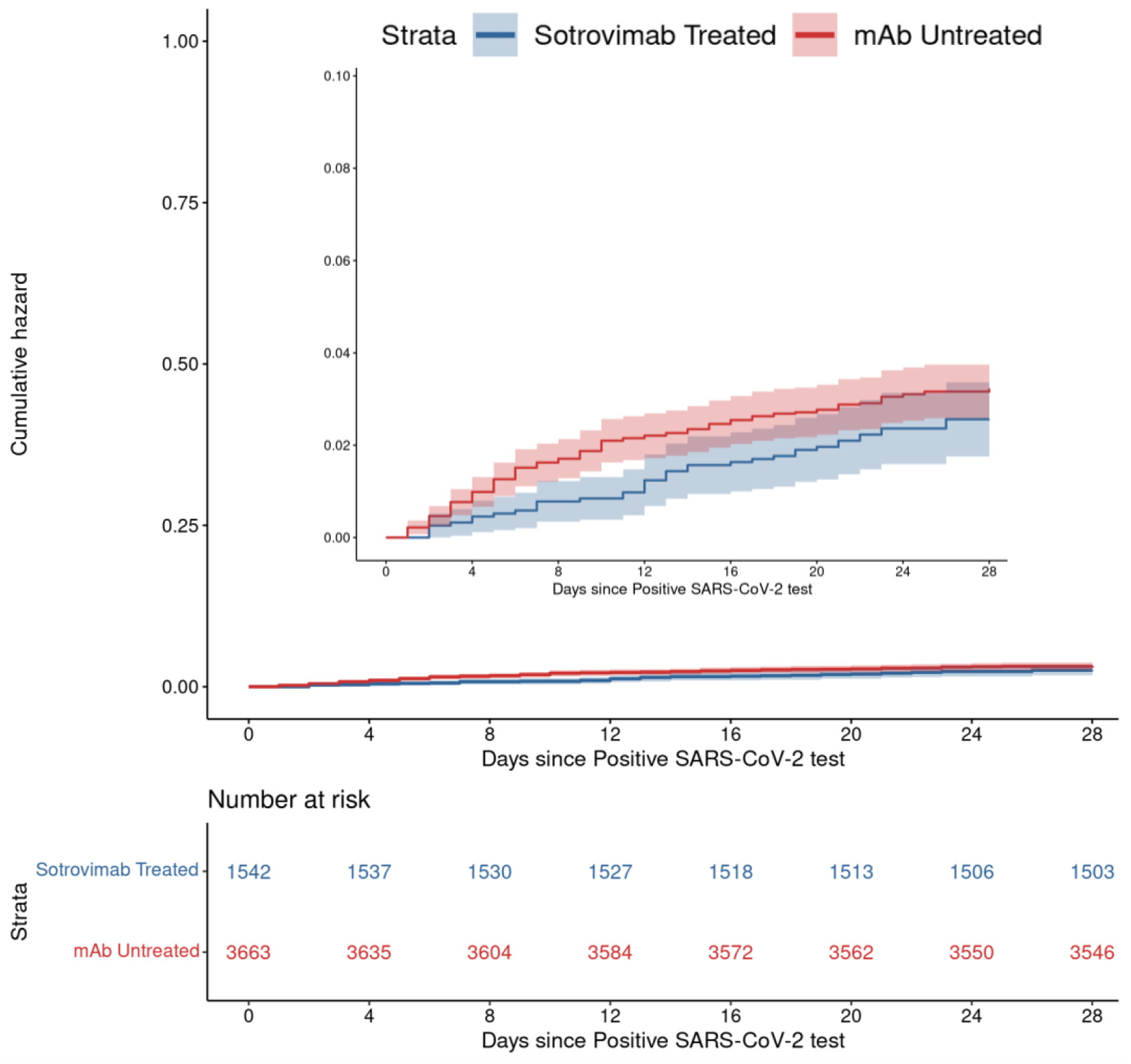
Cumulative Incidence Plots for All-Cause Hospitalization to Day 28 by Sotrovimab Treatment Status.

Rates of all-cause 28-day mortality were not statistically different, with 1 death in sotrovimab-treated group (0.1%) as compared to 7 deaths (0.2%) in the untreated group (aOR 0.62, 95% CI 0.07-2.78) (**Table 2**). ED visit rates were also similar between groups, 93 of 1542 (6.0%) in sotrovimab-treated, and 224 of 3663 (6.1%) in untreated (aOR 1.03, 95% CI 0.79-1.32).

### Severity of Hospitalization

Among hospitalized patients, 6 of 39 (15.4%) in the sotrovimab-treated group required high-flow nasal cannula (HFNC), non-invasive ventilation (NIV), invasive mechanical ventilation (IMV) or died in the hospital, compared to 33 of 116 (28.4%) in the untreated group (**Table 2**). The data also showed a higher proportion of sotrovimab-treated patients did not any require supplemental oxygen (35.9% vs. 17.2%). The average hospital length of stay (LOS) for sotrovimab patients was 5.2 (+/- 6.8) days in comparison to 7.3 (+/- 7.7) days in the untreated group. 4 of 39 (10.3%) sotrovimab-treated patients required ICU level of care, as compared to 20 of 116 (17.2%) untreated patients. Collectively, these data appear to suggest a lower severity of disease among hospitalized sotrovimab-treated patients, although the sample sizes were too low for valid statistical inference.

### Sotrovimab treatment effect in subgroups

During the Omicron phase, sotrovimab treatment was associated with a lower odds of 28-day hospitalization in older patients (age ≥ 65 years) as compared to no treatment (OR 0.52, 95% CI 0.30-0.92, interaction p=0.02). In addition, sotrovimab treatment trended towards a lower odds of 28-day hospitalization among immunocompromised (OR 0.63, 95% CI 0.38-1.04, interaction p=0.04) or having 2 or more comorbid conditions (OR 0.65, 95% CI 0.42 – 1.01, interaction p=0.007) as compared to no antiviral treatment (**Table 3**).

**Table 3.**
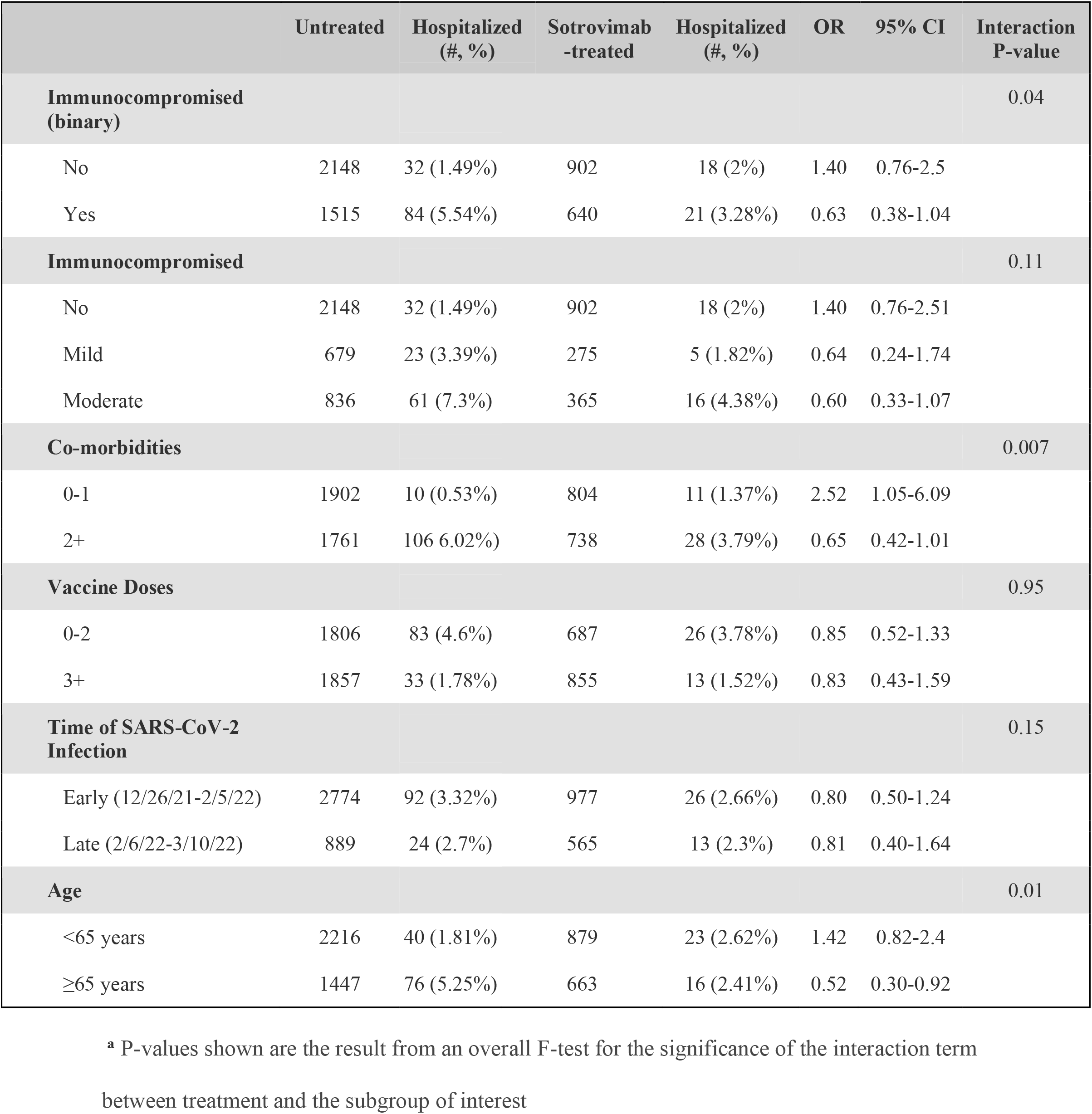
Treatment effects results among subgroups.

|  | Untreated | Hospitalized<br>(#, %) | Sotrovimab<br>-treated | Hospitalized<br>(#, %) | OR | 95% CI | Interaction<br>P-value |
| --- | --- | --- | --- | --- | --- | --- | --- |
| **Immunocompromised**<br>**(binary)** |  |  |  |  |  |  | 0.04 |
| No | 2148 | 32 (1.49%) | 902 | 18 (2%) | 1.40 | 0.76-2.5 |  |
| Yes | 1515 | 84 (5.54%) | 640 | 21 (3.28%) | 0.63 | 0.38-1.04 |  |
| **Immunocompromised** |  |  |  |  |  |  | 0.11 |
| No | 2148 | 32 (1.49%) | 902 | 18 (2%) | 1.40 | 0.76-2.51 |  |
| Mild | 679 | 23 (3.39%) | 275 | 5 (1.82%) | 0.64 | 0.24-1.74 |  |
| Moderate | 836 | 61 (7.3%) | 365 | 16 (4.38%) | 0.60 | 0.33-1.07 |  |
| **Co-morbidities** |  |  |  |  |  |  | 0.007 |
| 0-1 | 1902 | 10 (0.53%) | 804 | 11 (1.37%) | 2.52 | 1.05-6.09 |  |
| 2+ | 1761 | 106 (6.02%) | 738 | 28 (3.79%) | 0.65 | 0.42-1.01 |  |
| **Vaccine Doses** |  |  |  |  |  |  | 0.95 |
| 0-2 | 1806 | 83 (4.6%) | 687 | 26 (3.78%) | 0.85 | 0.52-1.33 |  |
| 3+ | 1857 | 33 (1.78%) | 855 | 13 (1.52%) | 0.83 | 0.43-1.59 |  |
| **Time of SARS-CoV-2**<br>**Infection** |  |  |  |  |  |  | 0.15 |
| Early (12/26/21-2/5/22) | 2774 | 92 (3.32%) | 977 | 26 (2.66%) | 0.80 | 0.50-1.24 |  |
| Late (2/6/22-3/10/22) | 889 | 24 (2.7%) | 565 | 13 (2.3%) | 0.81 | 0.40-1.64 |  |
| **Age** |  |  |  |  |  |  | 0.01 |
| <65 years | 2216 | 40 (1.81%) | 879 | 23 (2.62%) | 1.42 | 0.82-2.4 |  |
| ≥65 years | 1447 | 76 (5.25%) | 663 | 16 (2.41%) | 0.52 | 0.30-0.92 |  |

<sup>a</sup> P-values shown are the result from an overall F-test for the significance of the interaction term between treatment and the subgroup of interest

### Sotrovimab treatment effect in Omicron and Delta phases

Compared to treated patients during the Delta phase, treated patients in the Omicron phase were on average older, more white, more obese, more immunocompromised, had more comorbid conditions, and had been vaccinated with more doses (not shown). After propensity matching (**Appendix Figure 2**), these differences were no longer clinically meaningful or statistically different (SMDs <0.1). In the combined analysis across Omicron and Delta phases, the observed treatment OR for preventing 28-day hospitalization was higher during Omicron than during Delta predominance (OR 0.85 vs. 0.39, respectively; interaction p=0.053; **Appendix Table 4**).

## DISCUSSION

During a SARS-CoV-2 Omicron BA.1 variant-predominant phase in Colorado, sotrovimab was not associated with a lower incidence of the primary outcome of 28-day hospitalization. In addition, it was likely that sotrovimab treatment benefit observed during the Delta phase of the pandemic [2] was markedly attenuated during the Omicron BA.1 phase. However, the lower confidence boundary of 0.55 during the Omicron phase makes us interpret these results with caution. Notably, COVID severity metrics including all-cause mortality, hospital length of stay, as well as higher levels of respiratory support via HFNC, NIV, or invasive mechanical ventilation all trended in the direction of sotrovimab benefit, but was underpowered and did not reach statistical significance. Coupling these data with possible benefit from sotrovimab treatment among key subgroups (age ≥ 65 years, immunocompromised status, ≥ 2 comorbid conditions) that increase COVID severity, the results of our study are worthy of dissemination and for future reference as the COVID landscape continues to evolve.

Despite evaluating a cohort predominantly infected with Omicron BA.1 sublineages, our findings do not fully support observed sotrovimab neutralization of BA.1 variants *in vitro*,[23] though perhaps a lower sotrovimab neutralization potency against Omicron/BA.1 and Omicron/BA.1.1 as compared to ancestral strains and prior variants of concern made our findings more predictable.[13, 14] Further, with ineffective *in vitro* sotrovimab neutralization against Omicron BA.2 [13, 14] and among newer Omicron subvariants,[24] as well as a clinical observation that sotrovimab did not mitigate disease progression during a BA.2 Omicron dominant phase, [15] our findings do support the statements by the NIH guidelines committee [25] and FDA [26] that sotrovimab should not be recommended as a current outpatient treatment against COVID-19 among the general population of outpatients that meet EUA criteria. However, with a signal towards potential sotrovimab benefit in patients ≥ 65 years old, immunosuppressed, or with multiple comorbid conditions, some consideration should be given towards continued treatment in highest risk individuals depending on the availability of alternate treatments options, particularly if these observations continue to be made in other studies.[16]

Our results are of practical importance for policymakers and clinicians because there needs to be iterative data to support prioritization given shortages of mAb supplies and infusion capacity and other authorized antiviral treatments. As such, it is crucial to rapidly test real-world effectiveness of each treatment to mitigate hospitalization and mortality against each clinically relevant SARS-CoV-2 variant.[27]

### Limitations

This study has several limitations. Even though we used statewide data for mortality and vaccination status, hospitalizations were collected only within one single health system. In addition, this health system is geographically limited to one US state with relatively low racial and ethnic minority representation, though it serves both urban and rural populations through academic and community hospitals. If untreated patients were less likely to be seen in this health system, hence more likely to be hospitalized elsewhere, this may bias our results toward the null. We also relied on EHR data, including manual chart reviews, which may have missing or inaccurate information about the presence of chronic conditions.[28] These factors might have limited our ability to detect the impact of sotrovimab treatment.

We only collected 28-day hospitalization and mortality data, and therefore we cannot comment on sotrovimab effects over a longer phase after SARS-CoV-2 infection. However, our prior study would suggest that 28-day and 90-day data yield similar results with respect to hospitalization and mortality endpoints.[20] In this study, propensity scoring appropriately matched sotrovimab-treated and untreated patient groups across multiple variables, but unmeasured confounders may remain. Our EHR data does not contain information on SARS-CoV-2 variants at the patient level. However, during Colorado’s Delta phase more than 99% of sequenced SARS-CoV-2 was Delta variant and during Colorado’s Omicron phase it was more than 96% of Omicron BA.1.[17]

Finally, this study occurred while our health system’s sotrovimab distribution criteria changed due to implementation of austere measures, and as such, patients who received sotrovimab may have differed over the course of the study. We accounted for this by doing a subgroup analysis of early (12/26/21 – 2/5/22) and late (2/6/22 – 3/10/22) infection periods. Though we observed a similar sotrovimab effect in each period, it is notable that hospitalization rates among sotrovimab-treated and untreated groups appeared lower during the late period.

### Conclusion

This study of real-world data demonstrated sotrovimab treatment was not associated with reduced 28-day hospitalization among COVID-19 outpatients during the Omicron BA.1 variant phase. Outpatient sotrovimab treatment may still be beneficial in certain higher risk subgroups, and may reduce respiratory severity among those subsequently hospitalized, but larger cohorts are necessary to further examine these observations.

## Data Availability

All data produced in the present study are available upon reasonable request to the authors

## REFERENCES

1. Centers for Disease Control and Prevention. Science Brief: SARS-CoV-2 infection-induced and vaccine-induced immunity. Available at: https://www.cdc.gov/coronavirus/2019-ncov/science/science-briefs/vaccine-induced-immunity.html). Accessed April 25.

2. Aggarwal NR, Beaty LE, Bennett TD, et al. Real World Evidence of the Neutralizing Monoclonal Antibody Sotrovimab for Preventing Hospitalization and Mortality in COVID-19 Outpatients. J Infect Dis 2022.

3. Ganesh R, Pawlowski CF, O’Horo JC, et al. Intravenous bamlanivimab use associates with reduced hospitalization in high-risk patients with mild to moderate COVID-19. J Clin Invest 2021; 131(19).

4. Huang DT, McCreary EK, Bariola JR, et al. Effectiveness of casirivimab and imdevimab, and sotrovimab during Delta variant surge: a prospective cohort study and comparative effectiveness randomized trial. medRxiv 2021.

5. Jarrett M. Early Experience With Neutralizing Monoclonal Antibody Therapy For COVID-19. medRxiv 2021.

6. O’Horo JC, Challener DW, Speicher L, et al. Effectiveness of Monoclonal Antibodies in Preventing Severe COVID-19 With Emergence of the Delta Variant. Mayo Clin Proc 2022; 97(2): 327–32.

7. Razonable RR, Pawlowski C, O’Horo JC, et al. Casirivimab-Imdevimab treatment is associated with reduced rates of hospitalization among high-risk patients with mild to moderate coronavirus disease-19. EClinicalMedicine 2021; 40: 101102.

8. COVID-19 Treatment Guidelines Panel. Coronavirus Disease 2019 (COVID-19) treatment guidelines. Available at: https://www.covid19treatmentguidelines.nih.gov. Accessed April 27.

9. Dougan M, Nirula A, Azizad M, et al. Bamlanivimab plus Etesevimab in Mild or Moderate Covid-19. N Engl J Med 2021; 385(15): 1382–92.

10. Gupta A, Gonzalez-Rojas Y, Juarez E, et al. Effect of Sotrovimab on Hospitalization or Death Among High-risk Patients With Mild to Moderate COVID-19: A Randomized Clinical Trial. JAMA 2022.

11. Weinreich DM, Sivapalasingam S, Norton T, et al. REGN-COV2, a Neutralizing Antibody Cocktail, in Outpatients with Covid-19. N Engl J Med 2021; 384(3): 238–51.

12. Lynch HF, Caplan A, Furlong P, Bateman-House A. Helpful Lessons and Cautionary Tales: How Should COVID-19 Drug Development and Access Inform Approaches to Non-Pandemic Diseases? Am J Bioeth 2021; 21(12): 4–19.

13. Iketani S, Liu L, Guo Y, et al. Antibody evasion properties of SARS-CoV-2 Omicron sublineages. Nature 2022; 604(7906): 553–6.

14. Takashita E, Kinoshita N, Yamayoshi S, et al. Efficacy of Antiviral Agents against the SARS-CoV-2 Omicron Subvariant BA.2. N Engl J Med 2022; 386(15): 1475–7.

15. Zaqout A. Effectiveness of the neutralizing antibody sotrovimab among high-risk patients with mild to moderate SARS-CoV-2 in Qatar. medRxiv 2022.

16. Solera JT, Arbol BG, Alshahrani A, et al. Impact of Vaccination and Early Monoclonal Antibody Therapy on COVID-19 Outcomes in Organ Transplant Recipients During the Omicron Wave. Clin Infect Dis 2022.

17. Colorado Department of Public Health and Environment. Treatments for Covid-19. Available at: https://covid19.colorado.gov/for-coloradans/covid-19-treatments#collapse-accordion-40911-4. Accessed June 10.

18. ISPOR. About real-world evidence. December 2021. Available at: https://www.ispor.org/strategic-initiatives/real-world-evidence/about-real-world-evidence.

19. Angus DC. Optimizing the Trade-off Between Learning and Doing in a Pandemic. JAMA 2020; 323(19): 1895–6.

20. Wynia MK, Beaty LE, Bennett TD, et al. Real World Evidence of Neutralizing Monoclonal Antibodies for Preventing Hospitalization and Mortality in COVID-19 Outpatients. medRxiv 2022.

21. Georg Heinze MPaLJ. logistf: Firth’s Bias-Reduced Logistic Regression. R package version 1.24. 2020.

22. Team RC. R: a language and environment for statistical computing. Vienna: R Foundation for Statistical Computing, 2020.

23. Cameroni E, Saliba C, Bowen JE, et al. Broadly neutralizing antibodies overcome SARS-CoV-2 Omicron antigenic shift. bioRxiv 2021.

24. Yamasoba D, Kosugi Y, Kimura I, et al. Neutralisation sensitivity of SARS-CoV-2 omicron subvariants to therapeutic monoclonal antibodies. Lancet Infect Dis 2022.

25. National Institute of Health Covid treatment guidelines. Clinical spectrum of SARS-CoV-2 infection.. Available at: https://www.covid19treatmentguidelines.nih.gov/overview/clinical-spectrum. Accessed June 16.

26. FDA updates Sotrovimab emergency use authorization. Available at: https://www.fda.gov/drugs/drug-safety-and-availability/fda-updates-sotrovimab-emergency-use-authorization. Accessed 6/16/22.

27. Strategies to allocate scarce COVID-19 monoclonal antibody treatments to eligible patients examined in new rapid response to government. January 2021. Available at: https://www.nationalacademies.org/news/2021/01/strategies-to-allocate-scarce-covid-19-monoclonal-antibody-treatments-to-eligible-patients-examined-in-new-rapid-response-to-government). Accessed April 25.

28. Bennett TD, Moffitt RA, Hajagos JG, et al. Clinical Characterization and Prediction of Clinical Severity of SARS-CoV-2 Infection Among US Adults Using Data From the US National COVID Cohort Collaborative. JAMA Netw Open 2021; 4(7): e2116901.

