## Supplementary Material for "Change in Effectiveness of Sotrovimab for Preventing Hospitalization and Mortality in COVID-19 Outpatients During the Omicron Phase"

This Appendix has been provided by the authors to give readers additional information about the work.

### Appendix Figures

##### **Appendix Figure 1: Flow of Patients into the Primary Study Cohort**

89,005 subjects with a SARS CoV 2 positive test date on or before 3/10/22

Subject had a positive test prior to transition to Omicron variant (before 12/26/2021): N = 55,240

N = 33,765

N = 30,256

Propensity Matching

N = 1,542

N = 3,663

Subject had an order for molnupiravir (N = 62) or paxlovid (N = 356)

N = 30,247

N = 33,347

N = 32,870

Subsetting to mAb untreated (N = 31,187) and Sotrovimab treated (1,683) only

- Removed: treated with Evusheld within 10 days of the positive test (N = 379), bamlanivimab + etesevimab (N = 6), bebtelovimab (N = 2), casirivimab + imdevimab (N = 50), and inpatient remdesivir (40)

Missing both positive date and mAb administration date: N = 605

N = 32,265

Hospitalization Criteria: Removed N = 2,009

- Subject was admitted to hospital on the same day as their COVID + test (N = 141)
- Subject was already admitted to the hospital at the time of their COVID + test (N = 1,868)

Subject had more than 10 days between positive test date and mAb administration date: N = 9

##### Appendix Figure 2: Flow of patients into Delta Cohort

52,433 subjects with a SARS CoV 2 positive test date on or before 12/11/2021

N = 16,277

N = 9,463

N = 10,984

Subject had more than 10 days between positive test date and mAb administration date: N = 9

N = 9,457

Missing both positive date and mAb administration date: N = 948

N = 11,932

Subsetting to mAb untreated (N = 11,369) and Sotrovimab treated (563) only

Subject had a positive test prior to beginning of Delta cohort phase (before 10/01/2021): N = 36,156

Hospitalization Criteria: Removed N = 1,521

- Subject was admitted to hospital on the same day as their COVID + test (N = 109)
- Subject was already admitted to the hospital at the time of their COVID + test (N = 1,412)

N Untreated = 8,901

N Treated = 556

##### Appendix Tables:

**Appendix Table 1:**

| **Category** | **Medications** | **Conditions** |
| --- | --- | --- |
| Not Immunocompromised | No qualifying medications | No qualifying conditions |
| Mild  (Presence of either medication or condition if moderate/severe criteria not met) | - TNF-alpha inhibitors (infliximab, etanercept, golimumab, adalimumab) - Azathioprine alone - Mycophenolate alone - Systemic Prednisone - Systemic Methylprednisolone | - Elixhauser Rheumatic - Charlson or Elixhauser HIV without mention of AIDS |
| Moderate/Severe  (Presence of either medication or condition) | - Alemtuzumab - Azathioprine plus mTORi or calcineurin inhibitor - Belatacept - Eculizumab - Rituximab - Cyclophosphamide - Mycophenolate plus mTORi or calcineurin inhibitor - Thymoglobulin - Any Calcineurin inhibitor alone - Any mTORi alone - Any Chemotherapeutic Agent - Actinomycin - Alkylating Agent - Anthracenedione - Anthracycline - Anti-Metabolite - Anti-Microtubular - Aromatase Inhibitor - CDK Inhibitor - Cytotoxic - EZH2 Inhibitor - Hedgehog Inhibitor - Immunomodulatory Imide - Immunotherapy - Microtubule Inhibitor - mTOr Kinase Inhibitor - PI3K Inhibitor - Platinum-Based - Poly (ADP-Ribose) Polymerase Inhibitor - Proteasome Inhibitor - Targeted Monoclonal Antibody - Topoisomerase Inhibitor - Tyrosine Kinase Inhibitor - Vinca Alkaloid | - Charlson or Elixhauser HIV with AIDS based on CD4 count <200 cells/μL - Lymphoma - Cancer with Metastases - Tumor |

##### Appendix Table 2. Baseline Characteristics by Monoclonal Antibody Treatment Status for Full SARS-CoV-2 Positive Cohort, Prior to Propensity Matching

| **Characteristic** | **mAb-Treated**  **n=1663** | **mAb-Untreated n=28584** |
| --- | --- | --- |
| **Age Group *** |  |  |
| 18-44 years | 408 (24.5%) | 17326 (60.6%) |
| 45-64 years | 519 (31.2%) | 8012 (28.0%) |
| ≥65 years | 736 (44.3%) | 3246 (11.4%) |
| **Sex *** |  |  |
| Female | 979 (58.9%) | 16645 (58.2%) |
| **Race/Ethnicity *** |  |  |
| Non-Hispanic White | 1334 (80.2%) | 18594 (66.3%) |
| Hispanic, any race | 169 (10.2%) | 4565 (16.0%) |
| Non-Hispanic Black | 43 (2.6%) | 1600 (5.6%) |
| Other | 90 (5.4%) | 2320 (8.1%) |
| Missing | 27 (1.6%) | 1145 (4.0%) |
| **Insurance Status *** |  |  |
| Private/Commercial | 802 (48.2%) | 19465 (68.1%) |
| Medicare | 766 (46.1%) | 3309 (11.6%) |
| Medicaid | 52 (3.1%) | 3458 (12.1%) |
| None/Uninsured | 6 (0.4%) | 1113 (3.9%) |
| Other/Unknown | 37 (2.2%) | 1239 (4.3%) |
| **Immunocompromised *** |  |  |
| Mild | 286 (17.2%) | 2219 (7.8%) |
| Moderate / Severe | 387 (23.3%) | 1458 (5.1%) |
| **Obesity Status** |  |  |
| Yes | 508 (30.5%) | 4728 (16.5%) |
| Missing | 44 (2.6%) | 1709 (6.0%) |
| **Number of Other Comorbid Conditions *** |  |  |
| 0 | 423 (25.4%) | 16137 (56.5%) |
| 1 | 404 (24.3%) | 6230 (21.8%) |
| ≥2 | 792 (47.6%) | 4508 (15.8%) |
| Missing | 44 (2.6%) | 1709 (6.0%) |
| **Diabetes** |  |  |
| Yes | 393 (23.6%) | 2208 (7.7%) |
| Missing | 44 (2.6%) | 1709 (6.0%) |
| **Cardiovascular Disease** |  |  |
| Yes | 481 (28.9%) | 2226 (7.8%) |
| Missing | 44 (2.6%) | 1709 (6.0%) |
| **Pulmonary Disease** |  |  |
| Yes | 573 (34.5%) | 5383 (18.8%) |
| Missing | 44 (2.6%) | 1709 (6.0%) |
| **Renal Disease** |  |  |
| Yes | 286 (17.2%) | 1069 (3.7%) |
| Missing | 44 (2.6%) | 1709 (6.0%) |
| **Hypertension** |  |  |
| Yes | 816 (49.1%) | 5839 (20.4%) |
| Missing | 44 (2.6%) | 1709 (6.0%) |
| **Liver Disease** |  |  |
| Yes | 251 (15.1%) | 1544 (5.4%) |
| Missing | 44 (2.6%) | 1709 (6.0%) |
| **Number of Vaccinations *** |  |  |
| 0 | 355 (21.3%) | 9881 (34.6%) |
| 1 | 70 (4.2%) | 1691 (5.9%) |
| 2 | 303 (18.2%) | 9330 (32.6%) |
| 3+ | 935 (56.2%) | 7682 (26.9%) |
| **Days to mAb Administration** |  |  |
| N missing | 0 | 28584 |
| Mean (SD) | 2.95 (1.960) | NA |
| Range | 0.000 - 10.000 | NA |
| **Week *** |  |  |
| 12/26 - 1/1 | 5032 (17.6%) | 262 (15.8%) |
| 1/2 - 1/8 | 7255 (25.4%) | 108 (6.5%) |
| 1/9 - 1/15 | 6341 (22.2%) | 130 (7.8%) |
| 1/16 - 1/22 | 4435 (15.5%) | 172 (10.3%) |
| 1/23 - 1/29 | 2836 (9.9%) | 134 (8.1%) |
| 1/30 - 2/5 | 1248 (4.4%) | 195 (11.7%) |
| 2/6 - 2/12 | 637 (2.2%) | 180 (10.8%) |
| 2/13 - 2/19 | 328 (1.1%) | 124 (7.5%) |
| 2/20 - 2/26 | 241 (0.8%) | 122 (7.3%) |
| 2/27 - 3/5 | 134 (0.5%) | 137 (8.2%) |
| 3/6 – 3/10 | 97 (0.3%) | 99 (6.0%) |

Abbreviations: mAb, monoclonal antibody; SD, standard deviation; * variables used for propensity matching

##### Appendix Table 3. Full Model Results for 28-Day Hospitalization Primary Outcome

| Characteristic | Adjusted OR | 95% CI | P-value |
| --- | --- | --- | --- |
| Treatment Status |  |  |  |
| mAb-Untreated | Reference |  |  |
| Sotrovimab-Treated | 0.82 | 0.55, 1.19 | 0.294 |
| Age Group |  |  |  |
| 18-44 | Reference |  |  |
| 45-64 | 0.79 | 0.46, 1.36 | 0.382 |
| ≥65 | 1.68 | 1.02, 2.85 | 0.041 |
| Sex |  |  |  |
| Female | Reference |  |  |
| Male | 1.06 | 0.75, 1.49 | 0.74 |
| Race/Ethnicity |  |  |  |
| Non-Hispanic White | Reference |  |  |
| Hispanic, any race | 1.88 | 1.18, 2.90 | 0.009 |
| Non-Hispanic Black | 2.36 | 1.12, 4.54 | 0.026 |
| Other | 0.81 | 0.30, 1.79 | 0.631 |
| Insurance Status |  |  |  |
| Private/Commercial/Medicare | Reference |  |  |
| Medicaid | 1.39 | 0.57, 2.97 | 0.447 |
| Other (None/Uninsured/Unknown) | 1.86 | 0.59, 4.58 | 0.261 |
| Obesity Status |  |  |  |
| No | Reference |  |  |
| Yes | 1.52 | 1.08, 2.15 | 0.016 |
| Immunocompromised Status |  |  |  |
| No | Reference |  |  |
| Mild | 1.52 | 0.92, 2.46 | 0.097 |
| Moderate/Severe | 2.92 | 1.98, 4.35 | <0.001 |
| Number of Comorbid Conditions |  |  |  |
| 0 | Reference |  |  |
| 1 | 0.91 | 0.38, 2.17 | 0.821 |
| ≥2 | 4 | 2.06, 8.52 | <0.001 |
| Vaccination Status (# of doses) |  |  |  |
| 0 | Reference |  |  |
| 1 | 1.45 | 0.79, 2.56 | 0.225 |
| 2 | 0.62 | 0.39, 0.96 | 0.034 |
| ≥3 | 0.31 | 0.20, 0.46 | <0.001 |
| Time |  |  |  |
| One-week increase | 1.06 | 0.99, 1.13 | 0.1 |

Abbreviations: mAb, monoclonal antibody; OR, odds ratio; CI, confidence interval

|  | Delta Only Model | | | Omicron Only Model | | | Delta and Omicron Model | | |
| --- | --- | --- | --- | --- | --- | --- | --- | --- | --- |
|  | OR | 95% CI | P-value | OR | 95% CI | P-value | OR | 95% CI | P-value |
| Treatment Status |  |  |  |  |  |  |  |  |  |
| mAb-Untreated | Ref |  |  |  |  |  |  |  |  |
| Sotrovimab Treated | **0.4** | **(0.19, 0.77)** | **0.005** | **0.84** | **(0.52, 1.31)** | **0.453** | **0.39** | **(0.19, 0.74)** | **0.003** |
| Age in years |  |  |  |  |  |  |  |  |  |
| 18-54 | Ref |  |  |  |  |  |  |  |  |
| 45-64 | 1.71 | (0.77, 3.70) | 0.187 | 1.57 | (0.82, 2.98) | 0.169 | 1.62 | (0.98, 2.67) | 0.059 |
| ≥65 | 4.67 | (2.45, 9.36) | <0.001 | 3.05 | (1.81, 5.33) | <0.001 | 3.68 | (2.45, 5.66) | <0.001 |
| Sex |  |  |  |  |  |  |  |  |  |
| Female | Ref |  |  |  |  |  |  |  |  |
| Male | 1.35 | (0.81, 2.22) | 0.246 | 0.92 | (0.61, 1.37) | 0.677 | 1.04 | (0.76, 1.43) | 0.787 |
| Race/Ethnicity |  |  |  |  |  |  |  |  |  |
| Non-Hispanic White | Ref |  |  |  |  |  |  |  |  |
| Hispanic, all races | 1.25 | (0.50, 2.74) | 0.606 | 1.28 | (0.63, 2.37) | 0.478 | 1.22 | (0.71, 2.00) | 0.45 |
| Non-Hispanic Black | 3.02 | (1.13, 7.32) | 0.029 | 2.26 | (0.92, 4.90) | 0.074 | 2.44 | (1.27, 4.39) | 0.009 |
| Other | 0.97 | (0.33, 2.36) | 0.949 | 0.53 | (0.11, 1.60) | 0.295 | 0.71 | (0.30, 1.44) | 0.361 |
| Insurance Status |  |  |  |  |  |  |  |  |  |
| Private / Commercial / Medicare | Ref |  |  |  |  |  |  |  |  |
| Medicaid | 2.1 | (0.86, 4.73) | 0.102 | 1.85 | (0.65, 4.40) | 0.229 | 2.17 | (1.12, 3.93) | 0.023 |
| Other | 1.5 | (0.37, 4.53) | 0.534 | 4.21 | (1.58, 9.71) | 0.006 | 2.86 | (1.30, 5.67) | 0.011 |
| Obesity Status |  |  |  |  |  |  |  |  |  |
| No | Ref |  |  |  |  |  |  |  |  |
| Yes | 2.95 | (1.74, 5.01) | <0.001 | 1.46 | (0.96, 2.22) | 0.075 | 1.88 | (1.36, 2.61) | <0.001 |
| Immunocompromised |  |  |  |  |  |  |  |  |  |
| No | Ref |  |  |  |  |  |  |  |  |
| Mild | 1.67 | (0.80, 3.28) | 0.166 | 1.32 | (0.73, 2.31) | 0.35 | 1.35 | (0.86, 2.08) | 0.191 |
| Moderate/Severe | 1.25 | (0.61, 2.41) | 0.53 | 1.74 | (1.10, 2.77) | 0.018 | 1.59 | (1.09, 2.30) | 0.016 |
| No. of Comorbidities |  |  |  |  |  |  |  |  |  |
| 0 | Ref |  |  |  |  |  |  |  |  |
| 1 | 1.92 | (0.94, 4.04) | 0.073 | 0.99 | (0.41, 2.38) | 0.98 | 1.49 | (0.86, 2.63) | 0.153 |
| ≥2 | 2.44 | (1.17, 5.26) | 0.017 | 3.53 | (1.74, 7.74) | <0.001 | 3.17 | (1.90, 5.45) | <0.001 |
| Vaccine doses prior to infection |  |  |  |  |  |  |  |  |  |
| 0 | Ref |  |  |  |  |  |  |  |  |
| 1 | 0.21 | (0.04, 0.68) | 0.006 | 1.35 | (0.69, 2.50) | 0.37 | 0.77 | (0.42, 1.33) | 0.369 |
| 2 | 0.29 | (0.16, 0.51) | <0.001 | 0.59 | (0.36, 0.95) | 0.03 | 0.44 | (0.30, 0.63) | <0.001 |
| ≥3 | 0.17 | (0.03, 0.53) | 0.001 | 0.52 | (0.28, 0.92) | 0.025 | 0.38 | (0.22, 0.62) | <0.001 |
| Variant |  |  |  |  |  |  |  |  |  |
| Delta |  |  |  |  |  |  |  |  |  |
| Omicron |  |  |  |  |  |  | **0.55** | **(0.39, 0.79)** | **0.001** |
| Interaction |  |  |  |  |  |  |  |  |  |
| Treatment * Variant |  |  |  |  |  |  | **2.18** | **(0.99, 5.10)** | **0.053** |

##### Appendix Table 4. Delta Only and Omicron Only Models, and an Interaction Analysis

Abbreviations: mAb, monoclonal antibody; OR, odds ratio; CI, confidence interval; SD, standard deviation; other, none, uninsured, unknown;
